# Accounting for truncation artifacts in angiographic perfusion

**DOI:** 10.1101/2025.11.01.25339090

**Authors:** Kyle A Lyman, Ryan M Hebert, Charles C Matouk, Kevin N Sheth, Guido J Falcone, W. Taylor Kimberly, David Y Chung, Nils H Petersen, Ona Wu

**Affiliations:** Department of Neurology, Yale School of Medicine, New Haven, CT; Yale Center for Brain and Mind Health, Yale School of Medicine, New Haven, CT; Department of Neurosurgery, Yale School of Medicine, New Haven, CT; Department of Neurology, Massachusetts General Hospital, Harvard Medical School, Boston, MA; Athinoula A. Martinos Center for Biomedical Imaging, Massachusetts General Hospital, Harvard Medical School, Boston, MA

## Abstract

Unlocking perfusion metrics from routine digital subtraction angiography (DSA) could transform how neurovascular disease is managed. Truncation artifact, defined as premature termination of image acquisition, is a key source of error in CT and MR perfusion imaging and requires prolonged imaging. However, length requirements for deriving perfusion metrics from DSA remain undefined. This study investigates the minimum image acquisition length required for interpreting mean transit time (MTT) from DSA. We analyzed 55 outpatient angiograms performed for surveillance an average of 687 days (SD 541 days) after aneurysmal rupture. Truncation artifact was simulated by progressively shortening the angiography runs after bolus arrival. A gamma function was used to assess if extrapolation of the truncated data could approximate full length acquisitions. Extrapolation of the truncated datasets with the gamma function produced highly reliable estimates of the MTT with at least 6 seconds of post-bolus data. The fraction of pixels fit by the gamma function was more sensitive to truncation and ≥ 7 seconds of data was required to fit >90% of the pixels. These findings provide a framework for evaluating the adequacy of image acquisition time and lay the foundation for retrospective perfusion analysis using DSA.

## Introduction

Aneurysmal subarachnoid hemorrhage (aSAH) remains a major cause of morbidity and mortality^1–3^. In the absence of standardized monitoring protocols, management varies widely across centers, complicating the interpretation of clinical trials and therapeutic outcomes^4–6^. Techniques such as transcranial doppler, near-infrared spectroscopy, EEG, and invasive neuromonitoring are inconsistently available, underscoring the need for a universally accessible imaging biomarker to guide aSAH management.

Digital subtraction angiography (DSA) is routinely performed in all centers managing aSAH^7^. For decades, DSA interpretation has relied on subjective assessments of vessel caliber, filling patterns, and contrast transit times. These operator-dependent evaluations lack standardization to guide clinical decision making. However, DSA provides high temporal and spatial resolution data that are universally archived in DICOM format^8^ and making them ideal for quantitative analysis. Unlike qualitative review, which is limited to descriptive assessments, quantitative perfusion analysis can extract objective, reproducible metrics with defined physiological correlates. In our recent work, we demonstrated that block-circulant singular value decomposition (bcSVD^9^) can be applied to DSA time–intensity curves to compute the time-to-maximum of the residue function (Tmax)^10^. By deconvolving the signal-time curve of individual pixels with a single arterial input function (AIF), bcSVD provides a reliable estimate of perfusion parameters and is used in most commercial perfusion algorithms^9^.

Unlike CT and MR perfusion, which use three-dimensional voxels^9^, DSA-based perfusion is inherently two-dimensional, limiting direct assessment of cerebral blood flow (CBF) and cerebral blood volume (CBV). Mean transit time (MTT), defined as CBV/CBF^11^, is expressed in units of time without the need for an accurate assessment of the volume of tissue that each pixel represents. This physiologically meaningful parameter reflects vascular compliance and autoregulatory capacity^12^, hence measuring MTT represents an important goal for using DSA-based perfusion. Compared with Tmax, MTT is more sensitive to truncation artifact^13^, which occurs when acquisition ends before full venous contrast washout. Prior work in CT and MR perfusion has shown that truncation leads to systematic underestimation of MTT^13,14^. Yet, the temporal constraints necessary for reliable MTT estimation from conventional angiography have not been established.

We hypothesized that MTT could be accurately derived from conventional angiograms if sufficient post-bolus data were acquired. To test this, we analyzed long-duration outpatient angiography runs from patients with a history of aSAH, progressively truncating acquisitions to simulate shortened scans. We then evaluated whether model-based extrapolation could mitigate truncation effects.

## Methods

### Study Design

We retrospectively evaluated a population of adult (>18 years old) patients who had been admitted to Massachusetts General Hospital (MGH) from 6/2017 until 4/2023 for management of aneurysmal subarachnoid hemorrhage and who subsequently had outpatient conventional angiography performed for surveillance. DSA performed during these patient’s initial hospitalization for management of aSAH were studied in our prior publication^10^. The study was approved by the institutional review board of MGH with a waiver of informed consent and was conducted in accordance with the Declaration of Helsinki. To minimize confounding from acute vascular pathology, only surveillance angiograms interpreted as normal in the procedural report were included.

### Perfusion Analysis

Analyses were restricted to lateral projection films, and all angiogram frames were downsampled to 512 x 512 pixels for speed of processing. All data were upsampled to 4 Hz^10^. The unit of analysis was a single angiography ‘run’, defined as one injection of contrast with serial image acquisition spanning the arterial, capillary, and venous phases. For consistency and reproducibility, we followed a previously validated pipeline^10^. We began by using a rectangular region of interest placed over an extradural portion of the internal carotid artery (ICA) using DICOM metadata to determine the spatial scale in millimeters. The vessel diameter was calculated at each pixel and the concentration of intravascular iodinated contrast was estimated^15^ assuming cylindrical ICA geometry^16^.

### Cerebral Pixel Selection

To avoid large vascular structures, a large hemispheric region of interest (ROI) was manually placed over the lateral film and the 30% of pixels with the highest relative cerebral blood volume (rCBV) were removed^17^. An ellipsoid was then fit onto the hemispheric ROI to account for variations in head tilt and only pixels between 15° and 180° were included to capture territories supplied by the anterior and middle cerebral arteries. The remaining pixels defined a capillary map (CapMap) representing the parenchymal pixels used for calculation of MTT (**Figure 1**).

**Figure 1).**
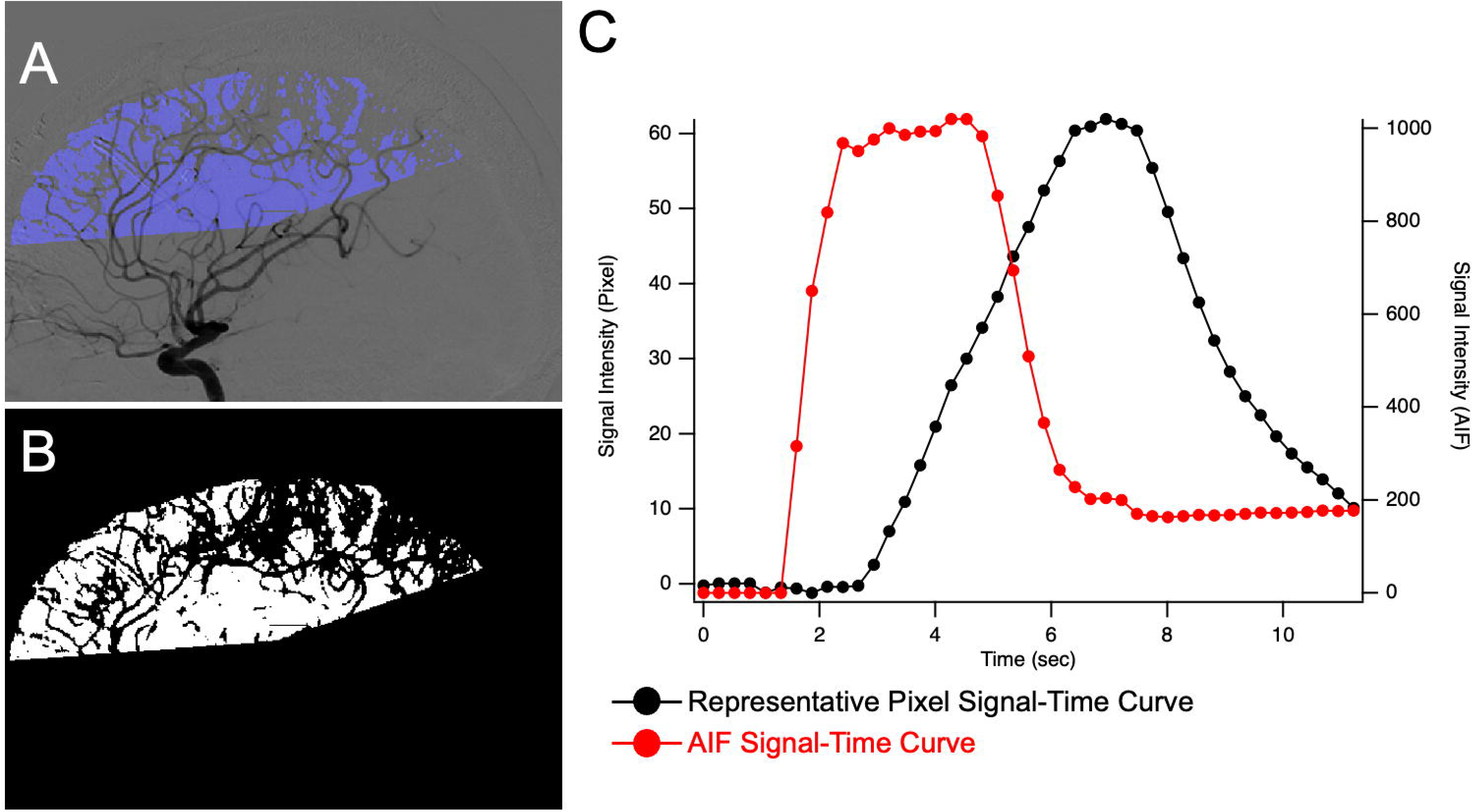
Nonvascular pixels in the region of the anterior cerebral artery (ACA) and middle cerebral artery (MCA) make up the CapMap. **A)** Schematic with angled region of interest (blue) superimposed onto the arterial phase of a representative angiogram. Note that the vascular structures are removed. **B)** Identical image to that shown in **(A)** with white pixels analyzed and black pixels representing those that were either vascular structures or outside of the region of interest. **C)** Signal-time curve for the arterial-input function (red) and a single pixel’s signal-time curve (black) after spatial averaging, as described in the text. AIF: Arterial input function.

### Deconvolution

Perfusion analysis was performed using block-circulant singular value decomposition with a threshold of 0.2^9^. Relative CBV (rCBV) was calculated as the integral of the deconvolved signal-time curve and relative cerebral blood flow (rCBF) defined as the maximum of the residue function^13^. MTT was defined as rCBV/rCBF by the central volume theorem^9^.

All analysis routines were performed using MATLAB (R2025a; Mathworks, Natick, MA). Figures were prepared using MATLAB and Igor Pro 9 (WaveMetrics, Portland, OR).

Axial perfusion modalities often consider the scan time after bolus arrival time to the cerebral vasculature^13,14^. We defined the bolus arrival time (BAT) as the time point prior to the maximum of the AIF when the second derivative of the AIF was maximal. All analyses of artifactually truncating the data are defined relative to the BAT.

Truncation artifact was simulated by progressively eliminating the final frames of each angiography run relative to the BAT (e.g., BAT+8 denotes removal of frames acquired more than 8 seconds after BAT), as shown in **Figure 2**.

**Figure 2).**
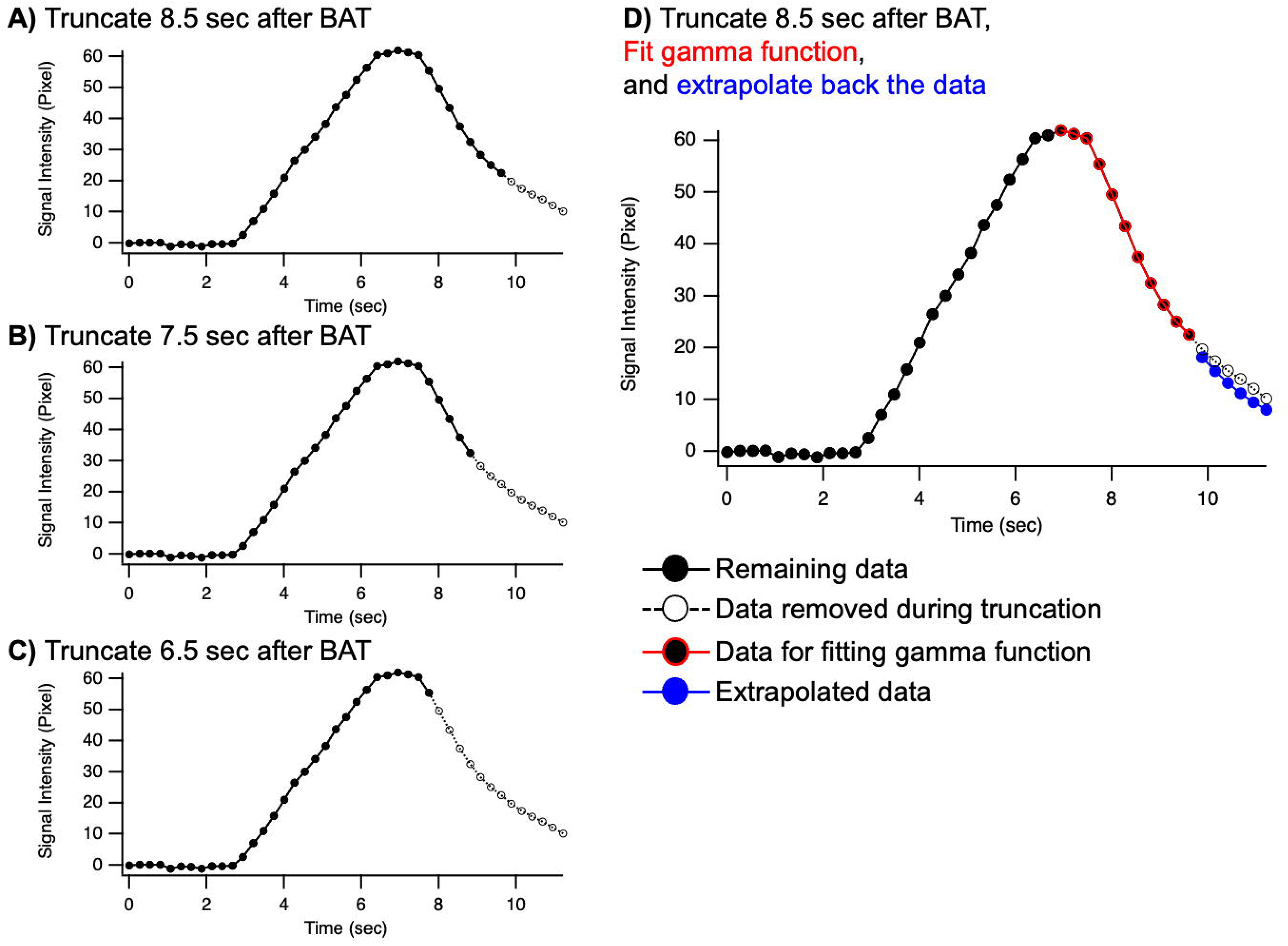
Truncation of a single pixel’s signal-time curve to simulate truncation artifact. **A-C)** The same pixel’s signal-time curve is shown with serial truncation of the data to 8.5 seconds after BAT (**A**), 7.5 seconds after BAT (**B**), and 6.5 seconds after BAT (**C**). In each case, the data that are used for subsequent analysis of truncation are shown in filled black circles with the original data in open circles connected by a dashed line. **D)** Truncated data can be extrapolated with the gamma function. Shown in panel **D** is the same data presented in panel **A** but with the datapoints used for fitting the gamma function in red (from the peak of the signal time curve until the truncation). In blue are the data points that are extrapolated using the gamma function fit to the red data points. The error term is calculated as the root mean squared error of the open circles (original data) compared with the blue data (extrapolated) and normalized by the mean of the open circles. BAT: Bolus Arrival Time

### Gamma Function Modeling and Extrapolation

To assess whether truncated data could be extrapolated, the descending portion of each pixel’s signal-time curve was modeled with a gamma function of the form^15,18^:

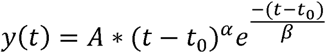

Nonlinear least-squares fitting was performed using MATLAB’s lsqcurvefit function, minimizing the sum of squared residuals between observed and predicted values. Data removed to simulate truncation were not used for fitting the gamma function (**Figure 2**). For each angiography run, the population-averaged CapMap signal-time curve was fit to derive initial parameters, which were then applied for pixel-level fitting. To improve robustness, each pixel’s signal-time curve was spatially averaged with neighboring pixels within a 3-pixel radius contained within the CapMap. Excluded vascular pixels were not used for averaging.

For each truncation length, pixel-level gamma fits were extrapolated to the full duration of the original angiography run (e.g., if the original angiography run lasted for 10 seconds after the BAT, the extrapolated data for the BAT+6 analysis included 4 seconds of extrapolated data) as shown in **Figure 2D**.

### Assessment of Gamma Fit

For each angiography run and truncation level, two measures were used to assess the gamma fitting procedure outlined above. First, we examined the proportion of pixels within the CapMap where the gamma function converged. Second, we examined the normalized root mean square error (RMSE) of the predicted versus observed signal, normalized by the mean signal intensity. We refer to this normalized RMSE term as R^-^MSE The median R^-^MSE value for each angiography run and truncation level was used for comparison (**Figure 3**).

**Figure 3).**
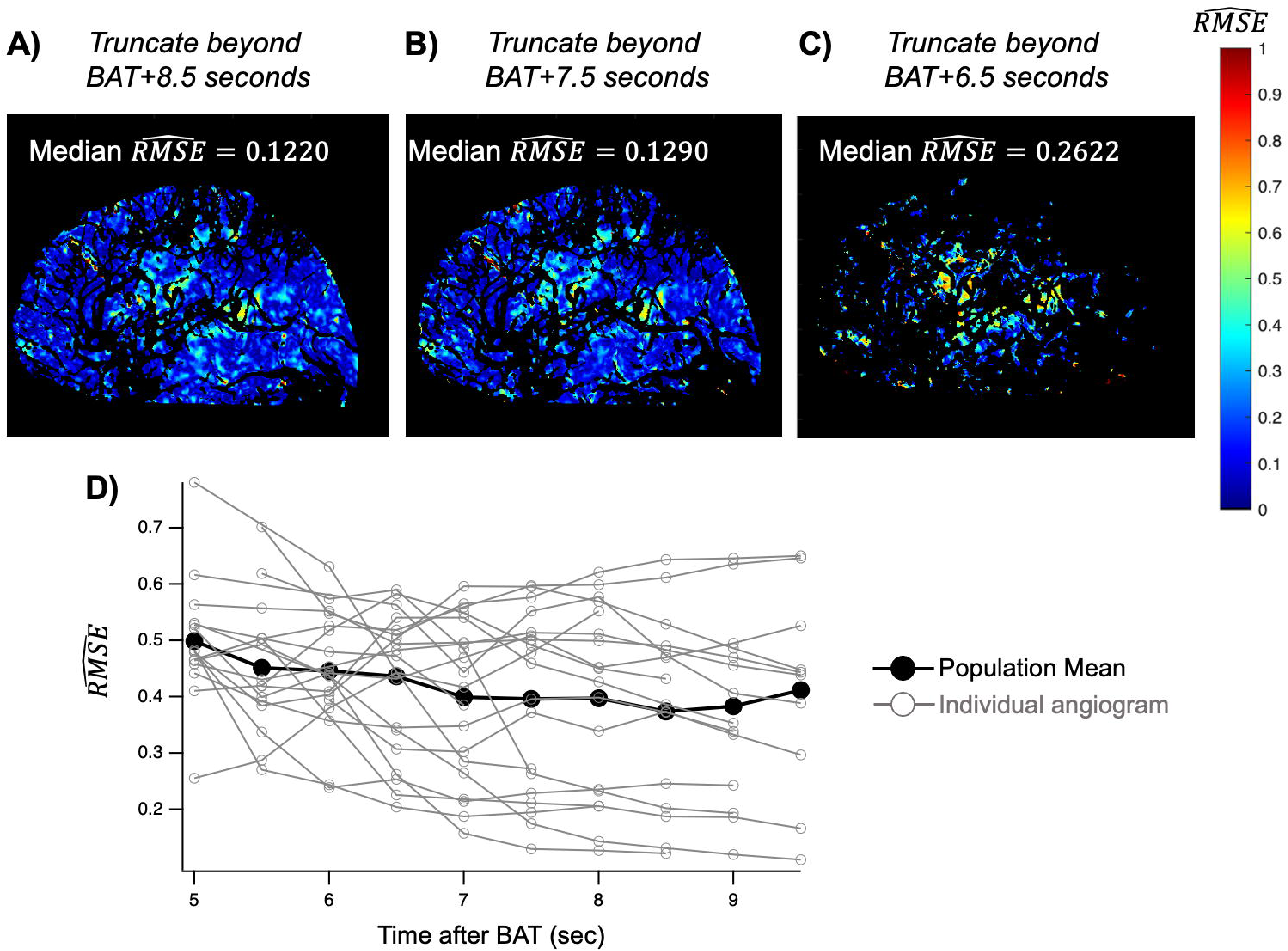
Progressive truncation of the data leads to fewer pixels fit by the gamma function and poor fitting of individual pixels. Panels **A-C** show the normalized error term maps associated with progressive truncation of the data. Note that in panel **C**, there are fewer pixels fit overall, and the pixels that are fit exhibit larger errors. **D)** Each angiography run’s median R^-^MSE value is shown as a function of the degree of truncation after BAT. BAT: Bolus Arrival Time

### MTT Estimation

After simulating truncation and extrapolating with the gamma function, the mean transit time from the extrapolated data (MTT_Extrap_) was calculated using bcSVD. Pixels where the gamma fit did not converge were not considered in the calculation of MTT_Extrap_. For each angiography run, MTT_Extrap_ was normalized to the original full-length MTT to examine the difference as a percent (100%*(MTT_Extrap_/MTT) and an absolute error (|MTT_Extrap_ - MTT|).

### Statistics

In the text of the manuscript, all values are presented as mean plus or minus the standard error of the mean. In the figures, error bars represent standard error of the mean.

## Results

### Bolus characteristics and description of the cohort

Demographic information for the cohort is presented in **Table 1**. Across the 98 angiography runs in the cohort, the average length of image acquisition was 9.22 ± 1.95 seconds (mean ± SD). The full width at half maximum of the arterial input function (AIF) was 2.01 ± 0.62 seconds and the bolus arrival time (BAT) of the AIF occurred 1.72 ± 0.88 seconds after imaging acquisition began. For comparison, the bolus arrival time in axial perfusion modalities (CTP, MRP) typically occupies ∼10 seconds of the initial portion of the scan and dispersion of the contrast leads to a full width at half maximum of the AIF of 10-15 seconds^14,19,20^. The length of image acquisition after the BAT was 7.49 ± 2.09 seconds.

**Table 1:**
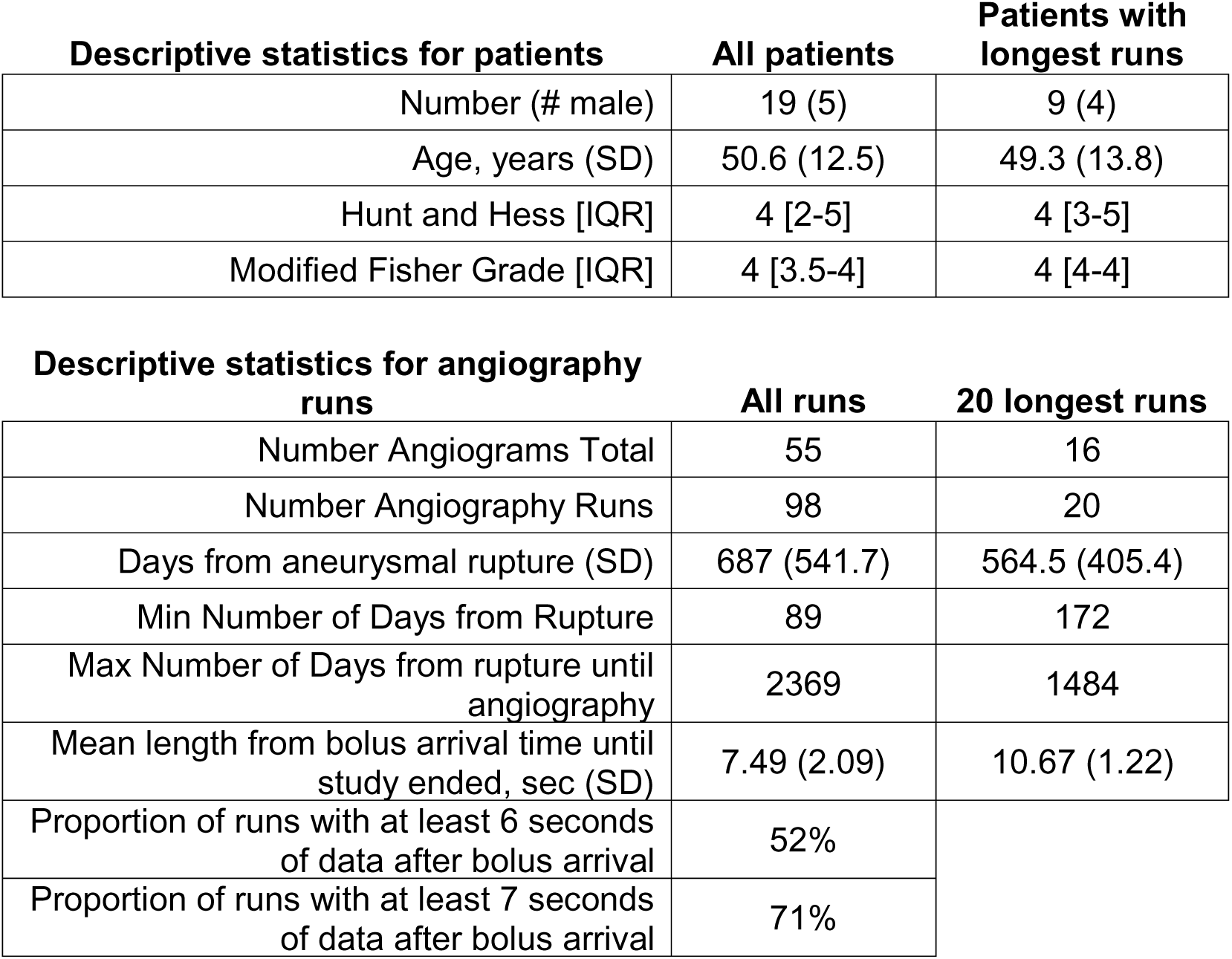
Patient demographics during their initial hospitalization for aneurysmal rupture.

To investigate the influence of truncation artifact, we chose the 20 angiography runs with the longest imaging acquisition after BAT (each at least 9.5 seconds). In this subset, the mean BAT was 1.23 ± 0.58 s and the duration of image acquisition after BAT of 10.67 ± 1.22 s.

### Effect of Truncation on Mean Transit Time (MTT)

As outlined in the methods section, we were specifically interested in a region of interest without vascular contamination and whose vascular supply was exclusively the internal carotid artery (ICA). The average MTT in this CapMap region across the 20 angiography runs was 2.79 ± 0.44 s (mean ± SD, **Figure 4**).

**Figure 4).**
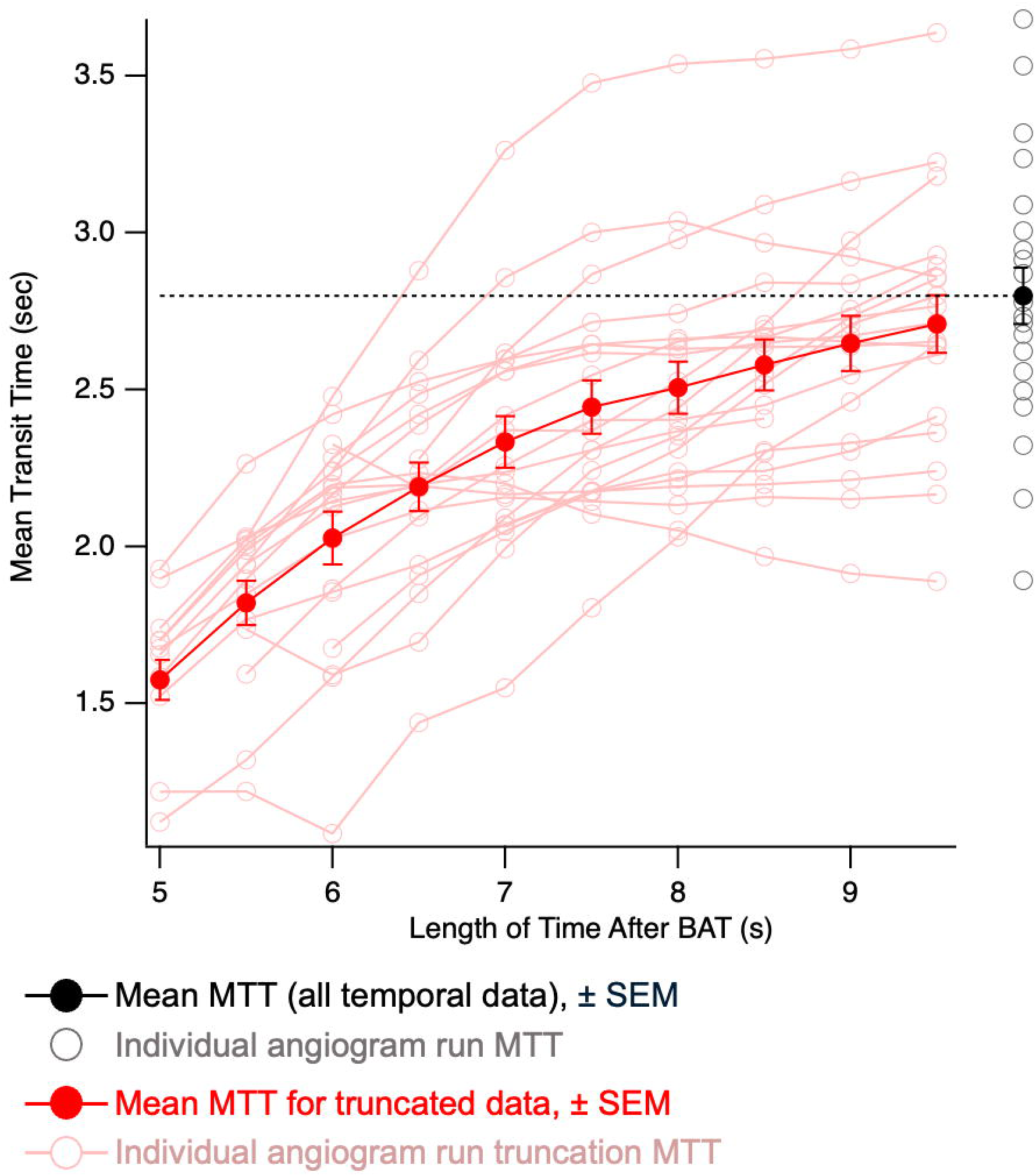
Truncation of the data leads to underestimation of the MTT. The filled black circle represents the mean MTT across angiography runs when no truncation was performed. Open black circles denote the MTT calculated from individual angiography runs without truncation. The dashed black line highlights the mean MTT from the angiography runs without truncation for reference. Filled red circles denote the mean MTT from the 20 angiography runs truncated to the length of acquisition time shown on the X axis. In pink, the calculated MTT from each angiography run is shown after truncating the data to the time point specified. MTT: Mean Transit Time. BAT: Bolus Arrival Time. SEM: Standard Error of the Mean

To evaluate the effect of incomplete angiography acquisitions, we progressively truncated the signal-time curves at successive intervals after the BAT (e.g., BAT+9.0s, BAT+8.5s, etc).

Truncation of the data led to underestimation of the MTT, with the degree of underestimation increasing as more data were removed. When data were truncated to 5 s after BAT, the calculated MTT decreased to less than 60% of the original value, highlighting the relevance of truncation artifact to accurately estimating MTT **(Figure 4/5)**.

### Gamma Extrapolation

Prior work has established that contrast kinetics can be accurately modeled with a gamma function (see Methods) hence we next investigated if the signal time curve from the truncated files could be extrapolated to improve the estimate of MTT. For each angiography run and truncation length, we first examined the number of pixels that converged within the CapMap. **Figure 6** shows the proportion of pixels that converged declines with increasing truncation. There was an apparent inflection point at 7 seconds of data after BAT, where 90.22 ± 9.9% of pixels converge.

We next examined the goodness of fit for each pixel. For a given angiography run, each pixel’s fit was compared with the non-truncated data to compare the extrapolation with the original result (**Figure 2D**, R^-^MSE is calculated comparing the blue extrapolated points with the original data, shown in open circles). This led to a family of R^-^MSE values for each angiography run and each truncation length as shown in **Figure 3**. Comparing the median R^-^MSE value for the 20 angiography runs across the different truncation lengths, we observed that the pixel-level error term appeared to stabilize at 7s of data after the BAT (**Figure 3D**).

### Accuracy of the Gamma-Extrapolated Signal for MTT

For each of the truncated angiography runs, we computed the extrapolated MTT (MTT_Extrap_) and normalized it to the original, non-truncated MTT calculated for that angiography run using the original dataset (**Figure 5**). MTT_Extrap_ using only the subset of pixels where the gamma function converged. Gamma extrapolation markedly improved the accuracy of MTT estimates compared to uncorrected truncation. With just 5 seconds of data after the BAT, the MTT_Extrap_ was 93.0 ± 7.3% (mean ± SD) of the original MTT value, and improved to 99.91 ± 1.67% (mean ± SD) when 7 seconds of data after the BAT were used.

**Figure 5).**
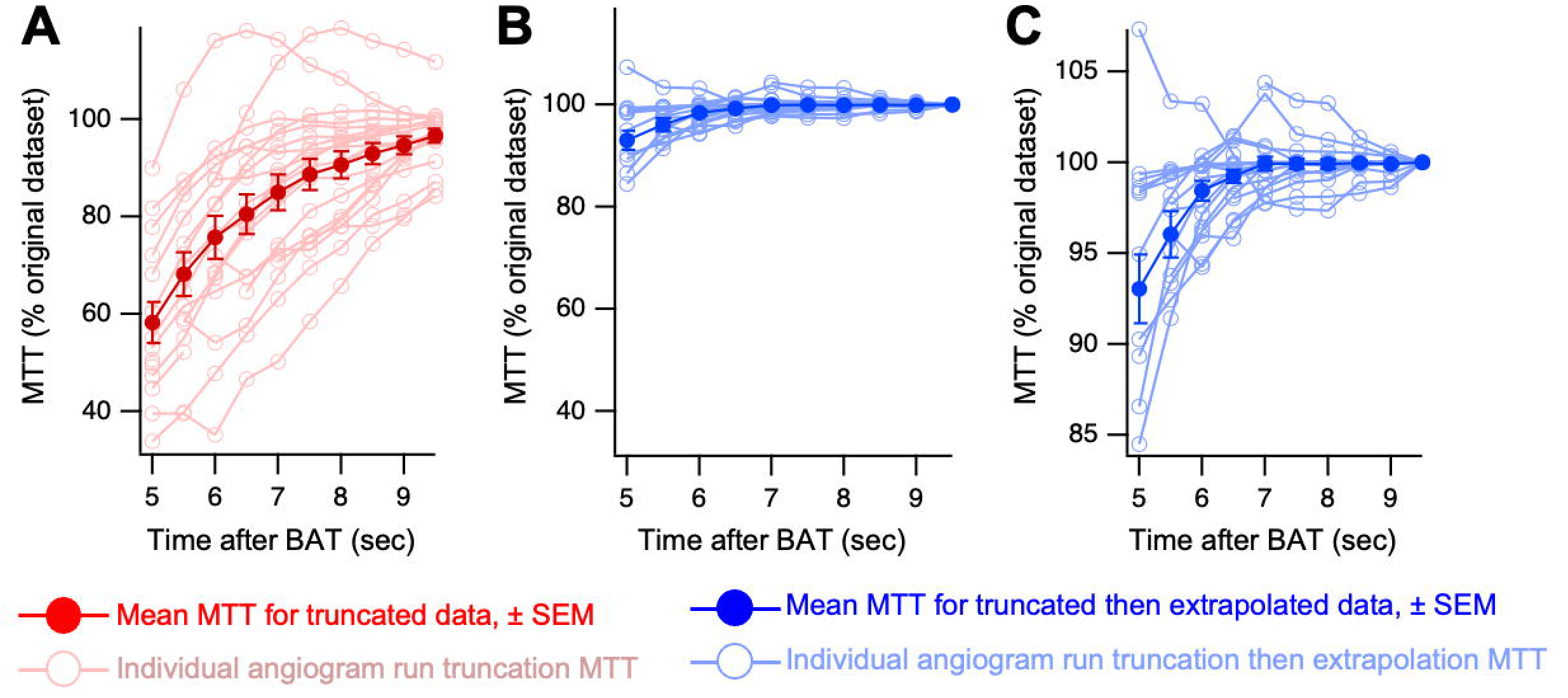
Extrapolation of the truncated data produces an accurate estimate of the MTT. **A)** The MTT from each of truncated datasets is plotted as a percentage of the MTT for that angiography run. Note that this is the same data presented in Figure 4 but normalized to each angiography run’s MTT when calculated from the original, non-truncated data. **B)** The MTT calculated from the extrapolated data (MTT_Extrap_) is plotted (blue) as a function of the degree of truncation followed by extrapolation. For example, the filled circle at Time after BAT = 6 indicates the effect of truncating the data to 6 seconds after BAT, extrapolating each angiography run back to the original length using the gamma function, and then calculating the MTT from the extrapolated dataset. Note that the Y axis is scaled identically in **A** and **B** for comparison. **C)** The same data from **B** are plotted with magnification of the Y axis for clarity. Even with severely truncated data, extrapolation of the signal-time curves for each angiography run leads to a reasonable estimate of the MTT. MTT: Mean Transit Time. BAT: Bolus Arrival Time. SEM: Standard Error of the Mean

**Figure 6).**
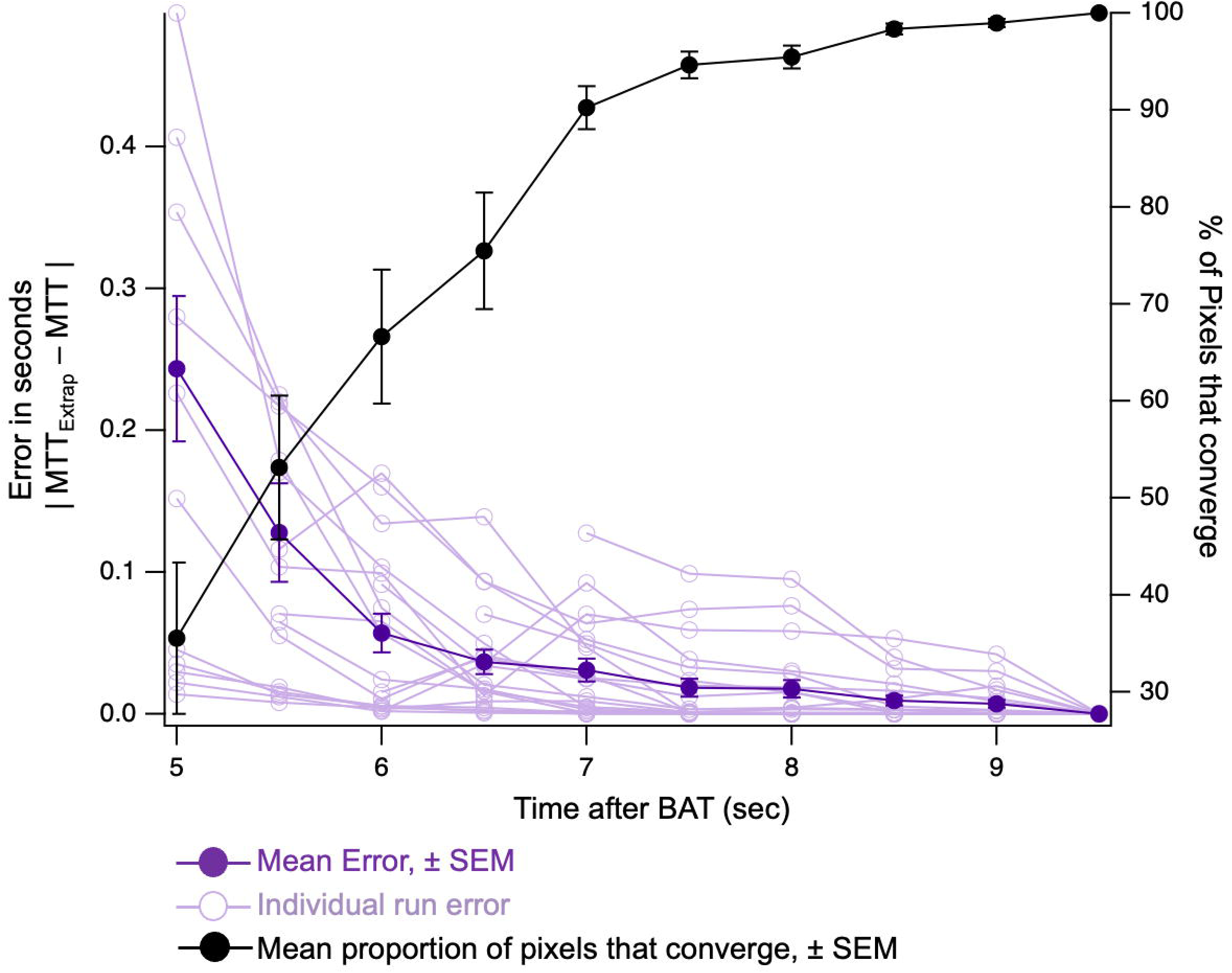
Pixel convergence provides a more sensitive readout of truncation artifact. In black, the mean proportion of pixels where the gamma function converges is plotted (right Y axis). When at least 7 seconds of data after the bolus arrival time are used, the gamma function converges for greater than 90% of pixels. In purple, the error term associated with each truncation is plotted, comparing the original, non-truncated MTT with the MTT derived from extrapolating the signal-time curves (MTT_Extrap_). While the gamma function converges for fewer pixels when there is significant truncation, the pixels that do converge produce an estimate of MTT that is accurate. The error term does not significantly increase until the data are truncated to less than 6 seconds of data after the bolus arrival. MTT: Mean Transit Time. BAT: Bolus Arrival Time. SEM: Standard Error of the Mean

To evaluate the performance of this extrapolation, we examined the absolute difference between MTT_Extrap_ and the true (non-truncated) MTT (**Figure 6**). Errors increased sharply when less than 6 seconds of data after BAT were used. Notably, while the fraction of fitted pixels decreased at shorter acquisition lengths, extrapolated MTT values remained highly accurate for datasets with at least 6 seconds after BAT. These results demonstrate that gamma-extrapolation can compensate for moderate truncation artifact in angiographic perfusion data.

## Discussion

In this report, we established a framework for evaluating the adequacy of angiographic acquisitions for estimating mean transit time (MTT) and provided quantitative estimates of MTT under physiologic conditions. Consistent with prior work on axial perfusion imaging^13,14^, serial truncation of data led to systematically shorter MTT values, underscoring the importance of truncation artifact in digital subtraction angiography (DSA)–based perfusion analysis. Extrapolating the signal–time curve using a gamma function model offered additional insight into data adequacy and recovery of physiologic MTT estimates.

The fraction of pixels that could be successfully fit by the gamma function emerged as the most sensitive indicator of truncation artifact. With 6 seconds of data after bolus arrival (BAT), the extrapolated MTT (MTT_Extrap_) approximated 98.4 ± 2.3% of the full-length MTT, but only 66.6 ± 29.3% of pixels demonstrated model convergence. Extending acquisition to 7 seconds increased the proportion of convergent pixels to 90.2 ± 9.9%, while maintaining near-perfect agreement with the original MTT (99.9 ± 1.7%). These findings suggest that the percentage of pixels achieving gamma fit convergence may serve as a practical, image-derived quality metric for identifying and mitigating truncation artifact in DSA-based perfusion studies.

The ability to extract quantitative perfusion metrics from DSA represents a paradigm shift from traditional angiographic interpretation. Whereas conventional angiographic reports describe findings using subjective terms (e.g. “mild delay” or “moderate vasospasm”), quantitative perfusion analysis provides precise, continuous variables that enable statistical comparison, longitudinal tracking, and threshold-based clinical decision-making. This quantification facilitates several critical capabilities absent in qualitative assessment: (1) objective detection of subtle perfusion abnormalities that may be visually imperceptible, (2) precise measurement of treatment response with numerical effect sizes, (3) establishment of evidence-based thresholds for intervention, and (4) harmonization of data across multiple centers for meta-analysis and clinical trial design.

To our knowledge, no prior study has reported quantitative MTT values derived from bcSVD analysis of conventional angiography in healthy physiologic conditions. To minimize confounders such as vascular pathology and anatomic variation, we analyzed the longest outpatient surveillance angiograms and limited assessment to cortical territories supplied by the internal carotid artery. This approach yielded a narrow MTT distribution (2.79 ± 0.44 s), in excellent agreement with values reported from MR perfusion (2.8–3.0 s)^21^ and within the expected physiologic range below the circulation time (4.7–5.2 s) reported in prior angiographic studies^22^. These results support the physiologic validity of MTT estimates derived from DSA perfusion.

The finding that only ∼7 seconds of imaging after contrast arrival are required for accurate MTT estimation stands in contrast to axial perfusion techniques, which typically require >60 s of acquisition to avoid truncation artifact^13,14^. This difference likely reflects the faster kinetics and reduced temporal dispersion associated with intra-arterial contrast administration. Together, these data suggest that DSA-based perfusion may provide a rapid, reliable means of assessing cerebrovascular physiology. Critically, because DSA is universally performed in the management of aneurysmal subarachnoid hemorrhage (aSAH), extracting quantitative perfusion metrics from these routine studies addresses the inconsistent availability of multimodal neuromonitoring. Unlike qualitative angiographic assessment, which cannot provide standardized physiologic measurements across institutions, quantitative MTT offers a universal biomarker that transforms existing clinical data into research-grade perfusion measurements.

This approach enables retrospective analysis of existing angiographic databases, prospective multicenter data harmonization, and establishment of objective therapeutic targets^24^. Future studies should correlate DSA-derived MTT with concurrent multimodal neuromonitoring and perfusion imaging modalities to establish clinical relevance and validate quantitative thresholds for clinical decision-making that cannot be derived from qualitative assessment alone.

## Limitations

Several limitations warrant consideration. First, in the setting of acute vascular pathology, such as large-vessel occlusion, MTT prolongation may necessitate longer angiographic acquisitions to capture the complete venous outflow phase, even when gamma extrapolation is applied. Evaluating the fraction of pixels achieving convergence and serially assessing truncation effects may provide a practical framework for verifying study adequacy. Second, because DSA-based perfusion is inherently two-dimensional, our approach cannot distinguish between gray and white matter MTT. Prior studies have demonstrated regional heterogeneity in perfusion parameters^21,23^, and future work incorporating structural parcellation or volumetric correction would be necessary to address this limitation.

## Conclusions

Perfusion metrics derived from routine conventional angiography offer a promising opportunity to harmonize physiologic data across institutions due to the ubiquity of DSA. We demonstrate that quantitative MTT can be reliably estimated from as little as 6 seconds of post-bolus imaging and that the proportion of gamma-fit convergent pixels serves as a sensitive quality metric for truncation artifact. Future work should extend this framework to pathologic states and validate angiographic perfusion parameters against established neuromonitoring and imaging standards.

## Data Availability

All data produced in the present study are available upon reasonable request to the authors

## Notes

### Competing Interest Statement

The authors have declared no competing interest.

### Funding Statement

This study did not receive any funding

### Author Declarations

Massachusetts General Hospital Institutional Review Board determined that this retrospective, observational study constitutes non-human research

